# Can the impact of childhood adiposity on disease risk be reversed? A Mendelian randomization study

**DOI:** 10.1101/19008011

**Authors:** Tom G. Richardson, Eleanor Sanderson, Benjamin Elsworth, Kate Tilling, George Davey Smith

## Abstract

**Objective:** To evaluate whether early life adiposity has an independent effect on later life disease risk or whether its influence is mediated by adulthood body mass index (BMI).

**Design:** Two-sample univariable and multivariable Mendelian randomization.

**Setting:** The UK Biobank (UKB) prospective cohort study and four large-scale genome-wide association study (GWAS) consortia.

**Participants:** 453,169 participants enrolled in the UKB and a combined total of over 700,000 individuals from different GWAS consortia.

**Exposures:** Measured BMI during adulthood (mean age: 56.5) and self-reported adiposity at age 10.

**Main outcome measures:** Coronary artery disease (CAD), type 2 diabetes (T2D), breast cancer and prostate cancer.

**Results:** Individuals with genetically predicted higher BMI in early life had increased odds of CAD (OR:1.49, 95% CI:1.33-1.68) and T2D (OR:2.32, 95% CI:1.76-3.05) based on univariable MR (UVMR) analyses. However, there was little evidence of a direct effect (i.e. not via adult BMI) based on multivariable MR (MVMR) estimates (CAD OR:1.02, 95% CI:0.86-1.22, T2D OR:1.16, 95% CI:0.74-1.82). In the MVMR analysis of breast cancer risk, there was strong evidence of a protective direct effect for early BMI (OR:0.59, 95% CI:0.50-0.71), although adult BMI did not appear to have a direct effect on this outcome (OR:1.08, 95% CI:0.93-1.27). Adding age of menarche as an additional exposure provided weak evidence of a total causal effect (UVMR OR:0.98, 95% CI:0.91-1.06) but strong evidence of a direct causal effect, independent of early and adult BMI (MVMR OR:0.90, 95% CI:0.85-0.95). Weak evidence of a causal effect was observed in the MVMR analysis of prostate cancer (early life BMI OR:1.06, 95% CI:0.81-1.40, adult BMI OR:0.87, 95% CI:0.70-1.08).

**Conclusions:** Our findings suggest that increased CAD and T2D risk attributed to early life adiposity can be mitigated if individuals reduce their weight in later life. However, having a low BMI during childhood may increase risk of breast cancer regardless of changes to weight in later life, with timing of puberty also putatively playing an important role.

## Introduction

Childhood obesity is now widely recognised as a global public health crisis, yet its prevalence continues to rise^1 2^. Having a high body mass index (BMI) in early life is thought to increase the risk of various health conditions, such as coronary artery disease (CAD)^3^, type 2 diabetes (T2D)^4^ and different types of cancer^5–7^. Whether an individual can reverse the impact of childhood obesity through lifestyle modifications is unclear, particularly given that individuals who are obese in early life tend to remain obese in adulthood^8^. This makes it challenging to discern whether early life adiposity has an independent and lasting influence on disease risk, or if its effect is entirely mediated by later life adiposity. If the latter is the case, then the potential adverse consequences of childhood obesity could be avoided by attaining and maintaining a healthy weight in adulthood.

Mendelian randomization (MR) is an approach that can help address such issues, by harnessing genetic variants as instrumental variables to infer causality amongst correlated traits^9 10^. As an individual’s genotype is established at zygote formation then genetic variation is robust to reverse causation, and confounding is considerably less evident than in conventional observational studies^11^. Furthermore, recent methods have been developed to disentangle whether several exposures influence an outcome along the same causal pathway or whether they have independent effects^12 13^. One of these developments is known as multivariable Mendelian randomization^12 14^.

We have undertaken an MR study to evaluate whether genetically predicted early life BMI has an effect on 4 disease outcomes that have been linked to childhood adiposity: CAD, T2D, breast cancer and prostate cancer^3 4 7 15^. Genetic instruments were identified by undertaking a genome-wide association study (GWAS) of 453,169 individuals from the UK Biobank study with measures of BMI in adulthood (mean age=56.5) who also reported their body size at age 10. This allowed us to conduct both univariable and multivariable MR analyses to discern whether the predicted causal influence of early life BMI has an independent effect on disease risk or whether it is mediated through later life BMI.

We postulate that, if early life BMI has a causal influence on disease risk, then there are likely 3 main categories that such effects could be grouped into after accounting for adult BMI in a multivariable framework (Figure 1). Firstly, the influence of early life BMI on disease risk may be mediated by later life BMI. As such, early life BMI only has an indirect effect on disease risk via adult BMI, which for example could be attributed to individuals with a high BMI at age 10 remaining overweight into adulthood (Figure 1-1). Secondly, early life BMI may have only an independent (i.e. direct) effect on disease risk, which is not mediated through adult BMI (Figure 1-2), or it may also be plausible that BMI at separate timepoints may both influence disease risk via alternate causal pathways (i.e. early life BMI has both a direct and an indirect effect on outcome, Figure 1-3). Additional descriptions of direct, indirect and total effects within a multivariable framework can be found in Supplementary Figure 1.

**Figure 1:**
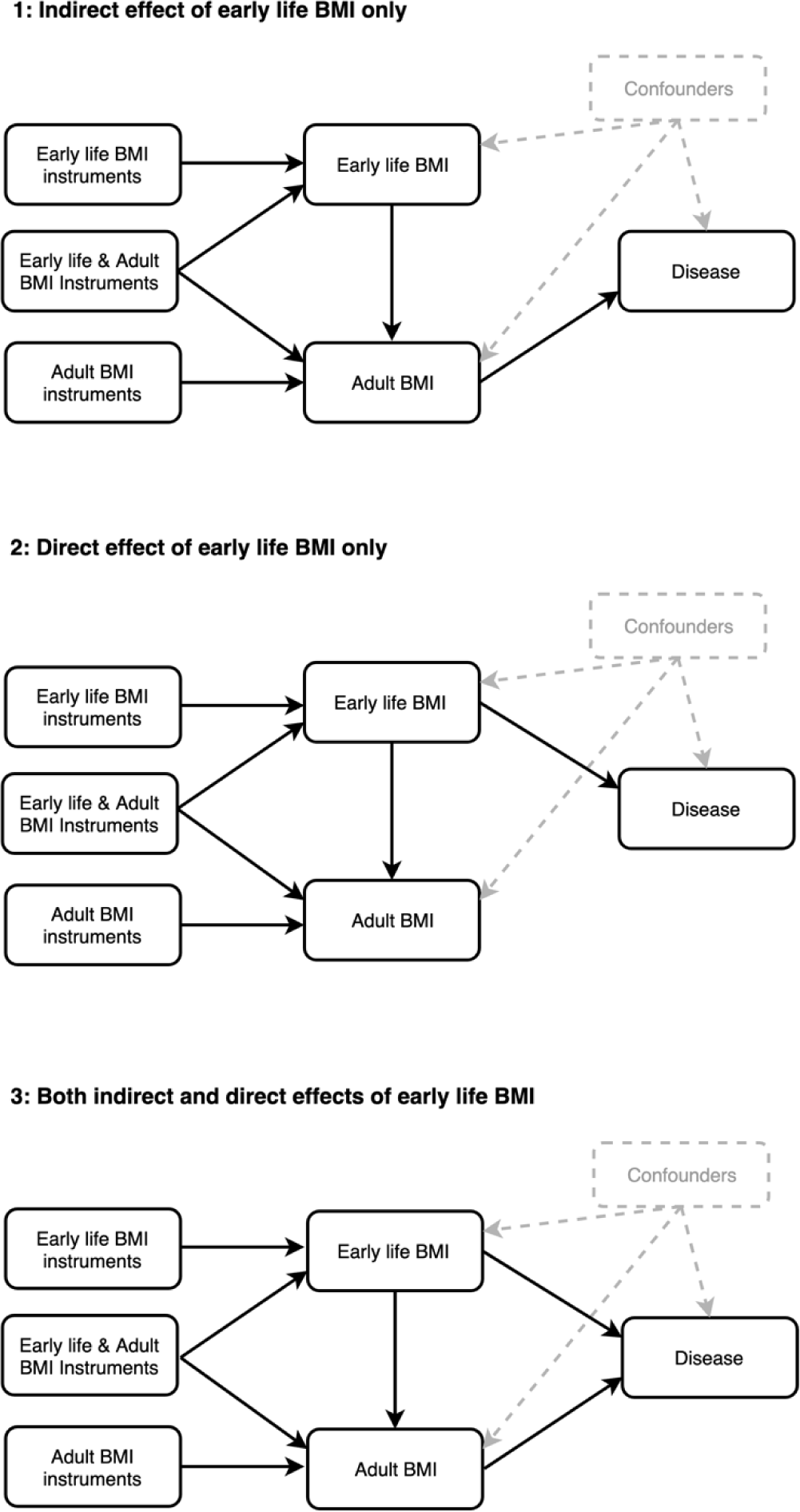
Directed acyclic graphs depicting three possible scenarios that could explain a causal effect between early life body mass index (BMI) and disease outcomes. 1) ‘Indirect effect of early life BMI only’ – early life BMI indirectly influences disease risk only via adult BMI 2) ‘Direct effect of early life BMI only’ – early life BMI has a direct effect on disease risk independent of adult BMI and 3) ‘Both indirect and direct effects of early life BMI’ –early life BMI exerts both direct and indirect effects on disease risk.

## Methods

### The UK Biobank and disease outcome datasets

Between 2006 to 2010 the UK Biobank study enrolled 500,000 individuals aged between 40 and 69 at baseline across 22 assessments centres in the United Kingdom^16^. Data were collected based on clinical examinations, assays of biological samples, detailed information regarding self-reported health characteristics and genome-wide genotyping^17^. BMI was derived using height (measured in whole centromeres) and weight (to the nearest 0.1kg) at baseline. Participants were also asked ‘When you were 10 years old, compared to average would you describe yourself as thinner, plumper or about average?’. Hereafter these measures are referred to as ‘adult BMI’ and ‘early life BMI’ respectively.

Only individuals with both BMI measures were included in analyses. Adult BMI was converted into a categorical variable with 3 groups based on the same proportions as the early life BMI variable (i.e. ‘thinner’, ‘plumper’ and ‘about average’). This was to ensure that both measures were as comparable as possible. Effect estimates from our results can be interpreted as the increase in odds conferred per additive change in BMI category. In total, 12,370,749 genetic variants in up to 463,005 individuals were available for analysis as described previously^18^. Effect estimates for genome-wide genetic variants on CAD^19^, T2D^20^, breast cancer^21^ and prostate cancer^22^ were obtained using findings from large-scale consortia which did not include data from the UK Biobank (Supplementary Table 1). Since earlier age at menarche is an established risk factor for breast cancer we also obtained summary statistics from a GWAS of age of menarche, which was used in additional analyses relating to this outcome^23^.

**Table 1:** Estimates from univariable and multivariable Mendelian randomization analysis assessing the effect of both predicted early life and adult body mass index on age at menarche. We also conducted a sensitivity analysis to investigate how both these exposures influence breast cancer risk when modelled with age at menarche as an additional third exposure. nSNPs – number of variants used as instrumental variables, Beta – effect estimate coefficient, - SE – standard error of the estimate, P – corresponding p-value.

| Outcome | Timepoint | nSNPs | Univariable analysis |  |  | Multivariable analysis |  |  |
| --- | --- | --- | --- | --- | --- | --- | --- | --- |
|  |  |  | Beta | SE | P | Beta | SE | P |
| *Age at menarche - Perry et al (2014)* | Age 10 | 102 | -0.93 | 0.14 | $1.42 \times 10^{-10}$ | -0.94 | 0.15 | $3.87 \times 10^{-10}$ |
|  | Adult | 142 | - | - | - | -0.03 | 0.14 | 0.82 |
| *Breast cancer - Michailidou et al (2017)* | Age 10 | 124 | -0.46 | 0.07 | $3.84 \times 10^{-11}$ | -0.64 | 0.10 | $1.57 \times 10^{-10}$ |
|  | Adult | 191 | - | - | - | 0.06 | 0.09 | 0.52 |
| | Age at menarche | 60 | -0.02 | 0.04 | 0.58 | -0.10 | 0.03 | $4.84 \times 10^{-03}$ |

### Statistical analysis

#### Identifying instruments by undertaking a genome-wide association study

The association between genetic variants across the human genome and both measures of BMI were assessed using the software BOLT-LMM^18 24^. This approach applies a Bayesian linear mixed model to account for both relatedness and population stratification. Age at baseline and type of genotyping array were added as covariates in the model. Analyses were undertaken 3 times, once in all eligible individuals after additionally adjusting for sex, and then in females and males separately. This allowed us to identify genetic variants which could be used as instrumental variables for outcomes based on a female-only population (i.e. breast cancer), a male-only population (i.e. prostate cancer) and mixed populations (i.e. CAD and T2D). Early life BMI analyses were additionally adjusted for month of birth as we hypothesized that an individual’s relative age within their school year may bias their reporting. Effect estimates from this analysis were used in the subsequent Mendelian randomization analyses.

Linkage disequilibrium (LD) clumping was undertaken using the software PLINK^25^ to identify independent variants from our GWAS. This was based on a threshold of r^2^ <0.001 using genotype data from European individuals (CEU) from phase 3 (version 5) of the 1000 genomes project as a reference panel^26^. For independent variants associated with either early life or adult BMI in our GWAS of all participants (based on P < 5 × 10^−08^), we compared their effect estimates between timepoints. This was undertaken by stacking observations (i.e. so that each individual had 2 BMI measurements) and then regressing the categorical BMI outcomes on each variant in turn adjusting for age, sex and time (i.e. early life or adulthood). This analysis was then repeated but also adjusting for the interaction between genetic variant and time. To account for multiple testing, we applied a Bonferroni correction to the p-values of the interaction terms (i.e. P < 0.05/number of individual variants assessed) as a heuristic to highlight genetic loci with the strongest evidence of an interaction with time. This analysis was undertaken to demonstrate that various genetic variants seemingly appear to have a stronger influence on BMI at different timepoints in the life courses.

We also undertook LD score regression^27^ to evaluate the genetic correlation between the two measures of BMI from our analysis with directly measured body mass index^28^ and childhood obesity^29^ using LD hub^30^. In doing so, we attempted to validate our early life measure of BMI by demonstrating that it is more strongly correlated with measured child obesity than adult BMI, given that is was derived using questionnaire data.

#### Mendelian randomization

Univariable MR analyses were conducted using the MR-Base platform^31^ to investigate total effect estimates between genetically predicted age 10 BMI and adult BMI individually with each disease outcome in turn. This was based on effect estimates using the inverse-variance weighted (IVW) method^32^. Effect estimates on early life BMI were used for the analysis with additional adjustment for month of birth. However, the “total” effect of adult BMI should be interpreted with caution. There is likely to be strong correlation between genetic determinants of early and adult BMI, and thus the total effect of adult BMI is likely to suffer from what will manifest as pleiotropy if there is also a direct effect of early BMI on the outcome. In the main manuscript we only discuss the total effect of early BMI, and direct effects of early and adult BMI, for this reason. A direct acyclic graph illustrating direct, indirect and total effects as investigated within a multivariable MR framework can be found in Supplementary Figure 1.

Multivariable MR analyses were undertaken to estimate the direct effect of early life BMI on each outcome in turn. Variants from the univariable analysis were used again here after undertaking further LD clumping to account for instrument correlation between the two sets. We also undertook a negative control analysis with early life and adult BMI as exposures in a multivariable framework and age at menarche as an outcome using findings from a GWAS published prior to the release of UK Biobank^23^. As depicted in Supplementary Figure 2, we hypothesized that early life BMI should only influence age at menarche directly in this framework as an individual’s adult BMI cannot influence timing of puberty. As such, direct and total effect derived from a univariable framework should be comparable for early life BMI on this outcome, whereas evidence of an indirect effect (i.e. via adult BMI) should be weak. Furthermore, we undertook a sensitivity analysis for the breast cancer outcome to discern whether timing of puberty may also play a role in disease risk. This involved repeating analyses on breast cancer with age at menarche as a third exposure (along with early life and adult BMI). Univariable and Multivariable MR analyses were also repeated using findings derived from ER-positive and ER-negative cases of breast cancer. Finally, we undertook simulated analyses to investigate how misclassification for early life BMI would affect our results (Supplementary Note 1).

All analyses were undertaken using R (version 3.5.1). Plots were created using the packages ‘ggplot2’^33^ and ‘metafor’^34^.

## Results

### A genome-wide association study of early life and adult body mass index

In a GWAS of 453,169 UK Biobank individuals, there were 295 and 557 independent associations detected with early life and adult BMI respectively based on conventional genome-wide corrections (i.e. P < 5 ×10^−08^) (Supplementary Tables 2 & 3). LD score regression analyses identified strong genetic correlation between our derived early life measure of BMI and a previous GWAS of childhood obesity (rg=0.85) (Supplementary Figure 3). In contrast, our adult BMI measure was considerably more strongly genetically correlated with a previous GWAS of adult BMI (rg=0.96) than to childhood obesity (rg=0.64). Evaluating the difference between the genetic variant associations at the two different timepoints suggested that 75 genetic variants had stronger effect estimates on BMI in adulthood compared to early life, whereas 23 genetic variants had a stronger effect in early life compared to adult BMI estimates (at P < 7.19 × 10^−05^ (i.e. 0.05/694 tests), Supplementary Table 4). Demonstrating that the magnitude of effect for these genetic variants differs with respect to early life and adult BMI suggests that they can be separated as two exposures in a multivariable framework.

Figure 2 illustrates findings from both GWAS using a bi-directional Manhattan plot (sometimes referred to as a ‘Miami plot’) where some examples of timepoint specific effects have been highlighted. Repeating our GWAS stratified by sex identified 135 and 215 variants which survived GWAS corrections (i.e. P < 5 × 10^−08^) in females only with early life and adult BMI respectively (n=246,511) (Supplementary Tables 5 & 6). In the male specific analysis, there were 69 genetic variants associated with early life BMI and 159 genetic variants associated with adult BMI (n=206,658) (Supplementary Tables 7 & 8). Repeating early life BMI analyses with additional adjustment for month of birth did not appear to materially influence overall findings (Supplementary Table 9-11).

**Figure 2:**
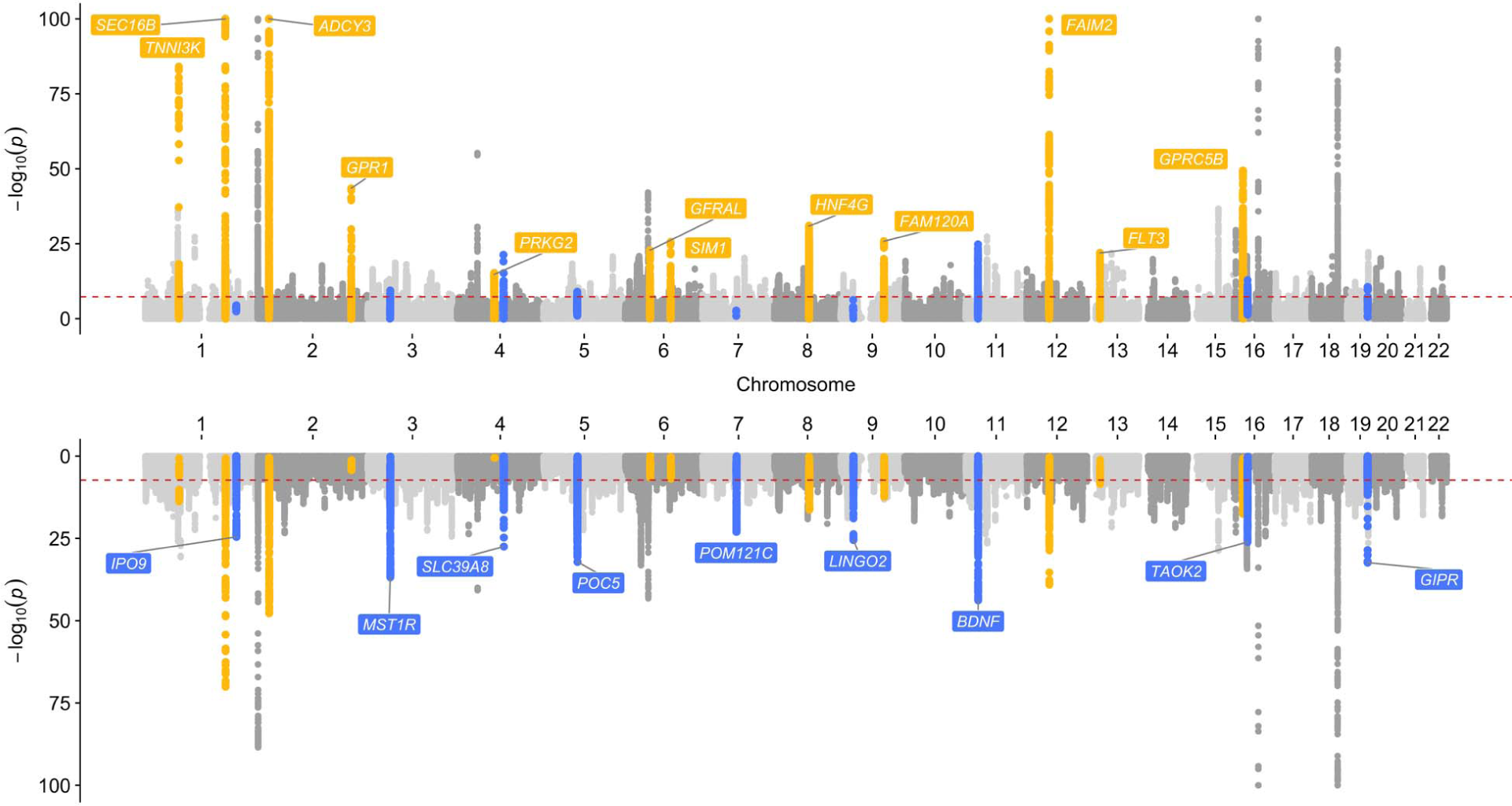
A bi-directional Manhattan plot to illustrate genetic variants across the genome associated with early life BMI (above) and adult BMI (below). Points highlighted into yellow highlight effects that provided stronger evidence of association with early life BMI, whereas points highlighted in blue were more strongly associated with adult BMI. These genetic loci have been annotated based on the nearest protein coding gene to the top associated variant.

### Univariable and multivariable Mendelian randomization analyses

Univariable analyses provided strong evidence that early life BMI is associated with CAD risk (UVMR OR:1.49, 95% CI:1.33-1.68). However, the direct effect of early life BMI (i.e. not mediated via adult BMI) was much smaller than the total effect (MVMR OR:1.02, 95% CI:0.86-1.22), whereas strong evidence of a direct effect was identified for adult BMI (OR:1.82, 95% CI:1.59-2.09). Similar findings were identified for analyses on T2D, as the magnitude of the direct causal effect for early life BMI was much smaller than the total effect when modelled simultaneously with adult BMI (early life BMI OR:1.16, 95% CI:0.74-1.82, adult BMI OR:2.80, 95% CI:1.89-4.15). Effect estimates from both univariable and multivariable MR analyses are illustrated in Figure 3 and a full list can also be found in Supplementary Table 12.

**Figure 3:**
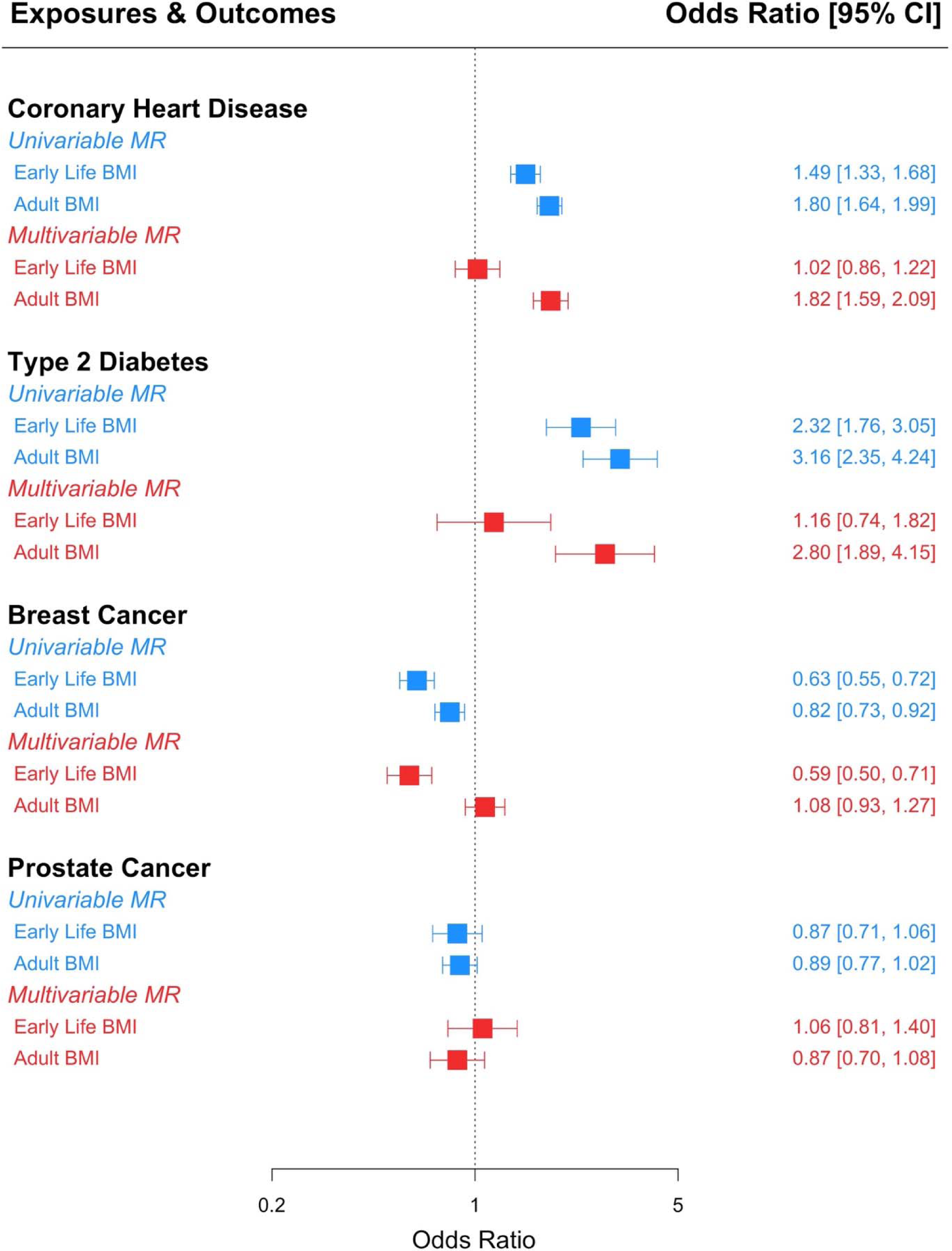
A forest plot illustrating direct and indirect effect for genetically predicted early life body mass index (BMI) on four different disease outcomes (coronary artery disease, type 2 diabetes, breast cancer and prostate cancer). Effects from the univariable and multivariable Mendelian randomization (MR) analyses are in blue and red respectively. Estimates are displayed as odds ratios with 95% confidence intervals (CIs).

The negative control analysis with age at menarche as an outcome provided strong evidence in the univariable analysis that early life BMI influences timing of puberty (Beta:-0.93, 95% CI:-0.66 to -1.20). Including adult BMI in a multivariable framework along with childhood BMI found that the direct effect estimate for childhood BMI was essentially unchanged, whilst adulthood BMI was not seen to influence age of menarche (Table 1). These findings reflect the scenario depicted in Supplementary Figure 2, suggesting that early life BMI can only effect timing of puberty directly and not indirectly via adult BMI (given that it occurs at an earlier timepoint in life course).

For breast cancer, there was strong evidence in the univariable analysis that higher genetically predicted early life BMI leads to reduced risk of this outcome (OR:0.63, 95% CI:0.55-0.72). Analysing both measures of BMI in a multivariable framework found that the indirect effect estimate via adult BMI (OR:1.08, 95% CI:0.93-1.27) was smaller in magnitude than the direct protective effect of early life BMI (OR:0.59, 95% CI:0.50-0.71). Similar findings were identified in the analysis of ER-positive and ER-negative cases of breast cancer (Supplementary Table 13).

Evaluating the relationship between genetically predicted age at menarche and breast cancer in a univariable analysis provided weak evidence of a total effect (OR:0.98, 95% CI:0.90-1.06). However, when modelled together with both early and adult BMI (i.e. all 3 exposures in the multivariable model) there was strong evidence that later age at menarche may lower the risk of breast cancer (OR:0.90, 95% CI:0.85-0.95). Effect estimates for predicted early life BMI had a larger effect in this multivariable analysis compared to the direct effect of adult BMI (Table 1). There was very weak evidence that early life BMI has a predicted causal effect on prostate cancer either directly (OR:1.06, 95% CI:0.81-1.40). or indirectly via adult BMI (OR:0.87, 95% CI:0.70-1.08).

Early life BMI is self-reported later in life and so is likely to be subject to misclassification which will not affect our measure of adult BMI. We therefore conducted a simulation to identify how such misclassification, affecting only one exposure, will influence the effect estimation in our multivariable MR analyses. Our results (see Supplementary Note 1) show that misclassification weakens the strength of association between the SNPs and our measure of early life BMI and so potentially biases the estimated effect of early life BMI in our multivariable MR analysis but does not introduce bias into the estimated effect between later life BMI and the outcomes. The results also show even with misclassification the effect of early life BMI in the multivariable MR estimation is not biased when it has a null effect on the outcome.

## Discussion

### Interpretation of findings

In this study we have examined the influence of early life adiposity on disease risk and whether this putative causal effect occurs independently (“direct effect”) or via the same causal pathway (“indirect effect”) as later life BMI. Our univariable MR analysis suggested that genetically predicted early life BMI increases risk of coronary artery disease and type 2 diabetes. However, when analysed together with adult BMI in a multivariable framework the direct effect estimates for early life BMI were of considerably reduced and essentially null magnitude in comparison to the estimates of the total effects, suggesting that its influence on these outcomes is mediated by later life BMI (as portrayed in the ‘Indirect effect of early life BMI only’ diagram in Figure 1). These findings imply that observed associations between early life obesity and increased coronary artery disease and type 2 diabetes risk are likely attributed to individuals with a high BMI during childhood remaining overweight into later life. This suggests that a window of opportunity exists to mitigate the detrimental impact of early life BMI on risk of these disease outcomes^35 36^. This corroborates findings from some previous research suggesting that there is no persistent influence of childhood obesity on risk of type 2 diabetes and cardiovascular disease, unless adiposity is sustained^37 38^. Furthermore, these findings highlight the importance taking into account adult BMI in order to assess whether childhood adiposity has a direct or persistent effect on disease risk over the life course^39 40^.

Our results also provide strong evidence that early life BMI has a protective effect on breast cancer risk as has been previously reported by both observational and MR studies which have not previously taken into account adult BMI^41–44^. This was identified both as a total and direct effect using univariable and multivariable MR analyses respectively, suggesting that the effect of early life BMI may persist into later life regardless of any interventions that influence BMI variation. Furthermore, reported protective effect estimates between later life BMI and breast cancer risk may be attributed to effects from childhood. We note however that this effect requires further investigation in subpopulations of pre- and post-menopausal breast cancer patients from large-scale GWAS. This is particularly crucial given observational evidence suggesting that higher BMI increases the risk of invasive breast cancer in post-menopausal women^45^.

As an additional validation of our multivariable framework, we undertook a negative control analysis to investigate the effect of early life BMI on at menarche in both a univariable and multivariable framework accounting for adult BMI. We found that estimates were consistent from both analyses for early life BMI, whereas the effect estimates for adult BMI in the multivariable analyses had a much smaller magnitude of effect in comparison. This provides a powerful proof of concept for our analytical framework, given that BMI can only influence timing of puberty in early life whereas adult BMI cannot as it occurs earlier in the life course.

We also undertook an additional analysis to investigate timing of puberty with respect to the putative predictive effect of early life BMI on breast cancer risk. To do this, we incorporated age at menarche as an additional exposure in our multivariable MR analysis. Our findings corroborate similar analyses undertaken using a multivariable framework, which suggests that later age at menarche has a protective effect on breast cancer risk but only when accounting for early life BMI^46^. However, there is strong evidence from the literature of a relationship between timing of puberty and later life BMI^47 48^. Notably, our results build upon previous findings by modelling age 10 and adult BMI as two separate exposures, due to the direct effect of age 10 BMI providing strong evidence of a protective effect when accounting for age at menarche. Similar observational findings have been reported recently in premenopausal women after adjusting for age at menarche^49^. Developing insight into the underlying biological mechanisms explaining this effect may therefore have important therapeutic implications.

We were unable to support previous evidence of an effect between genetically predicted BMI and prostate cancer risk^50 51^. This relationship is notoriously complex due to the effect of obesity on various hormones in males, such as an inverse relationship with prostate specific antigen^52^ and a positive relationship with estrogen concentrations^53^. This outcome is therefore worth revisiting using forthcoming findings from large-scale GWAS of prostate cancer when they become available, using this study’s analytical framework. A greater number of genetic instruments in males only is also likely to improve power for future endeavours. Evaluating the influence of pubertal development on this outcome would also be a worthwhile undertaking^54^.

We used univariable Mendelian randomization to estimate total effects of early BMI, and multivariable Mendelian randomization to estimate direct effects of early and adult BMI. In theory, the univariable estimate of the effect of adult BMI (the total effect) and the multivariable estimate (the direct effect) should be equal. However, if some genetic variants cause both early and adult BMI (albeit with different effect sizes), and early BMI has a direct effect on the outcome, then this will potentially generate pleiotropy in the univariable estimate of adult BMI. This can be seen in the univariable estimate of the effect of adult BMI on breast cancer, which is quite different to the multivariable estimate, probably due to the direct effect of early BMI on breast cancer. For CAD, where there is no direct effect of early BMI on the outcome, then the univariable and multivariable effects of adult BMI are the same. This demonstrates one of the issues with Mendelian randomization of time-varying exposures – the univariable analyses cannot identify critical or sensitive periods of exposure, but only an effect of a difference in the cumulative lifetime exposure^55 56^. Moreover, As outlined in all the diagrams depicted in Figure 1, we assume that childhood BMI has an effect on adulthood BMI in all scenarios. Therefore, individuals who (for example) lower their BMI between childhood and adulthood may effectively break this mediated causal chain and reduce their elevated risk of diseases such as coronary heart disease or type 2 diabetes.

### Strengths and limitations of this study

The key strengths of our investigation include the large number of individuals from the UK Biobank study with measures of both early and later life adiposity (n=453,169). This sample size is unprecedented compared to previous endeavours in the field, although we note that a caveat to this is having to rely on questionnaire-based data for the age 10 variable. As such there will likely be additional measurement error when analysing this exposure. The statistical power in our GWAS therefore comes at the expense of precision, although despite this we were able to recapitulate evidence of association between genetic variants and early life BMI measurements in the literature^57 58^.

Harnessing large-scale summary statistics from GWAS using two-sample MR is another strength of this study. This circumvents the necessity of having both our exposures and outcomes measured in the same sample. Moreover, along with using genetic variants to mitigate the influence of confounding and reverse causation, a multivariable MR framework has enabled us to investigate the independent effect of early life BMI, which is extremely challenging in an observational setting. Furthermore, although we have focused on 5 outcomes in this study, future work can adopt a similar approach to us for any outcomes of interest for which there are accessible GWAS summary statistics.

Our univariable analysis used all the SNPs associated with each exposure. Although there are variations in the SNPs associated with each of early life and adult BMI there is also considerable overlap in the SNPs associated with each timepoint. Our univariable MR analyses therefore capture pleiotropic effects of the SNPs through the other time point as well as, for childhood BMI, the effect mediated by adult BMI. This is a limitation of our univariable MR analyses but highlights the importance of the multivariable MR to disentangle the direct effects of child and adult BMI.

A limitation that should be taken into consideration when interpreting our results is that it is possible that participants from the UK Biobank may have also contributed to the large-scale GWAS whose results were used in this study. Overlapping exposure and outcome samples in a two-sample MR analysis may result in overfitting, although currently there is no way to robustly discern whether this is the case unless anonymous identifiers between the UK Biobank and GWAS consortium can be linked together^59^.

## Conclusions

Using multivariable Mendelian randomization, we have provided strong evidence suggesting that early life adiposity has a causal influence on breast cancer risk that acts independently of later life body mass index. Conversely, our results suggest that the positive association between early life adiposity and both coronary artery disease and type 2 diabetes is likely due to individuals with a high body mass index remaining overweight in later life. Our approach therefore yields insight into the mechanistic pathway between early life risk factors such as body mass index and disease outcomes. Furthermore, it provides scope to differentiate between effects where the conferred risk may not be reversable by achieving and maintain a healthy body mass index in adulthood.

### What is already known on this topic?

- Childhood obesity is known to have a detrimental impact on various health conditions and disease risk in later life.
- Body mass index in early life has been associated with increased odds of coronary artery disease and type 2 diabetes observationally, but whether this effect is immutable or whether lifestyle modifications can help mitigate it is unclear.
- The influence of early life body mass index on risk of other diseases, such as breast cancer and prostate cancer, has been previously reported but the causal relationships underlying these observations are complex and require further evaluation.

### What this study adds?

- The influence of early life body mass index on odds of coronary artery disease and type 2 diabetes was predominantly mediated through later life adiposity, suggesting that individuals can reduce the impact of any conferred risk by reducing their relative weight as they enter adolescence.
- Body mass index at age 10 had a protective effect on breast cancer risk which is independent of later life adiposity, although timing of puberty likely also plays a causal role in this relationship.
- Although in this study we have focused on cardiometabolic and cancer outcomes, the analytical approach we have used has considerable potential to investigate how risk factors trajectories across life influence different types of health outcomes.

## Data Availability

All data analysed in our study was made available from the UK Biobank study. Summary statistics will be made publicly available via the MR-Base platform.

## Acknowledgements

We are extremely grateful to the participants of the UK Biobank study and to the GWAS consortia (CARDIoGRAMplusC4D, DIAGRAM, BCAC and PRACTICAL) who made their summary statistics publicly available for the benefit of this study. This research was conducted at the NIHR Biomedical Research Centre at the University Hospitals Bristol NHS Foundation Trust and the University of Bristol. The views expressed in this publication are those of the author(s) and not necessarily those of the NHS, the National Institute for Health Research or the Department of Health. UK Biobank data was analysed under application 15825. UK Biobank has received ethical approval from the UK National Health Service’s National Research Ethics Service (ref 11/NW/0382).

